# Analysis of meteorological conditions and prediction of epidemic trend of 2019-nCoV infection in 2020

**DOI:** 10.1101/2020.02.13.20022715

**Authors:** Jin Bu, Dong-Dong Peng, Hui Xiao, Qian Yue, Yan Han, Yu Lin, Gang Hu, Jing Chen

## Abstract

**Objective:** To investigate the meteorological condition for incidence and spread of 2019-nCoV infection, to predict the epidemiology of the infectious disease, and to provide a scientific basis for prevention and control measures against the new disease.

**Methods:** The meteorological factors during the outbreak period of the novel coronavirus pneumonia in Wuhan in 2019 were collected and analyzed, and were confirmed with those of Severe Acute Respiratory Syndrome (SARS) in China in 2003. Data of patients infected with 2019-nCoV and SARS coronavirus were collected from WHO website and other public sources.

**Results:** This study found that the suitable temperature range for 2019-nCoV survival is (13-24 °C), among which 19°C lasting about 60 days is conducive to the spread between the vector and humans; the humidity range is 50%-80%, of which about 75% humidity is conducive to the survival of the coronavirus; the suitable precipitation range is below 30 mm/month. Cold air and continuous low temperature over one week are helpful for the elimination of the virus. The prediction results show that with the approach of spring, the temperature in north China gradually rises, and the coronavirus spreads to middle and high latitudes along the temperature line of 13-19 °C. The population of new coronavirus infections is concentrated in Beijing, Tianjin, Hebei, Jiangsu, Zhejiang, Shanghai and other urban agglomerations. Starting from May 2020, the Beijing-Tianjin-Hebei urban agglomeration, the Central China Zhengzhou-Wuhan urban agglomeration, the eastern Jiangsu-Zhejiang-Shanghai urban agglomeration, and the southern Pearl River Delta urban agglomeration are all under a high temperature above 24 °C, which is not conducive to the survival and reproduction of coronaviruses, so the epidemic is expected to end.

**Conclusions:** A wide range of continuous warm and dry weather is conducive to the survival of 2019-nCoV. The coming of spring, in addition to the original Wuhan-Zhengzhou urban agglomeration in central China, means that the prevention and control measures in big cities located in mid-latitude should be strengthened, especially the monitoring of transportation hubs. The Pearl River Delta urban agglomeration is a concentrated area of population in south China, with a faster temperature rise than those in mid-high latitudes, and thus the prevention in this area should be prioritized. From a global perspective, cities with a mean temperature below 24 °C are all high-risk cities for 2019-nCoV transmission before June.

## Introduction

Since Dec 15, 2019, the Chinese city of Wuhan has reported an outbreak of atypical pneumonia caused by the 2019 novel coronavirus (2019-nCoV)^1^. On January 14, 2020, the World Health Organization named the pathogen of this pneumonia as 2019 new coronavirus (2019-nCoV). As of February 4, 2020, the number of cases in mainland China has increased to a total of 20,492 confirmed cases and 23,214 suspected cases, causing significant negative impact both at home and abroad. As this coronavirus appears for the first time and is highly contagious, it poses a great challenge to diagnosis and prevention and control.

Similarly, from November 2002 to June 2003, there was another outbreak of coronavirus-caused pneumonia in China, known as severe acute respiratory syndrome (SARS), which eventually caused 5327 clinically diagnosed cases and 343 death ^2^ Studies have confirmed that the SARS outbreaks were significantly inverse associated with the temperature and its variations^3-5^. Based on the biological homology of the pathogens of two outbreaks of atypical pneumonia^6^, the present study intends to analyze the meteorological factors of the occurrence and transmission of new coronavirus pneumonia in 2019 with reference to the 2003 SARS meteorological data, and to summarize the meteorological characteristics of disease occurrence, and predict the development and regression of 2019-nCoV-caused pneumonia, with a view to provide basis for the prevention and control of pneumonia caused by 2019-nCoV.

## Material and Methods

The meteorological data of Wuhan from 2019-2020 and the meteorological data of Guangzhou from 1999-2003 were collected from Guangdong Meteorological Observation Data Center; the China national mean temperature data were collected from the National Climate Center of China Meteorological Administration; Global temperature data were collected from the USA National Center for Environmental Forecasting (NOAA, https://www.esrl.noaa.gov/psd/data/gridded/data.ghcncams.html). Data of patients infected cases by 2019-nCoV and SARS coronavirus were collected from WHO website and other public sources.

## Results

### 1. Meteorological conditions of onset of 2019-nCoV outbreak

The outbreak of atypical pneumonia caused by the 2019 novel coronavirus (2019-nCoV) has been reported in Wuhan since December 15. Considering the virus reproduction and disease incubation, we chose the period from Oct 1 to Dec 15 as subject for meteorological analysis.

In Oct, 2019, Wuhan entered winter with a stable winter climate, and the comparison of climates in Wuhan between 2019 and same period in previous years was showed in **Table 1**. The mean temperature in Wuhan from October to December in 2019 was 14.86 °C, which was 1.57 °C higher than that in 2015 and 0.64 °C higher than that in 2018. As shown in **Table 2**, there were 6 cooling processes in the winter of 2019, however, none lasted for more than 7 days. The impact of cooling process on the mean temperature was not significant.

**Table 1.** Comparison of climatic conditions from October 1 to December 15, 2019 and the same period of previous years in Wuhan.

| Parameters | 2015 | 2016 | 2017 | 2018 | 2019 |
| --- | --- | --- | --- | --- | --- |
| Days | 76 | 76 | 76 | 76 | 76 |
| Mean temperature (°C) | 13.29 | 13.51 | 12.82 | 14.22 | 14.86 |
| Mean humidity (%) | 83.92 | 83.12 | 80.90 | 78.26 | 75.26 |
| Total precipitation (mm) | 220.40 | 194.80 | 95.20 | 121.00 | 74.00 |

**Table 2.** Abnormal weather process in Wuhan before the outbreak of 2019-nCoV pneumonia

| Parameters | 2015 | 2016 | 2017 | 2018 | 2019 |
| --- | --- | --- | --- | --- | --- |
| Cooling process | 4 | 11 | 12 | / | 6 |
| Number of cooling process lasting more than 7 days | 3 times | 4 times | 5 times | Freezing days<br>Lasting about 40 days | None (only one time lasting 5 days) |
| Maximum temperature reduction in 24 hours (°C) | 7.52 | 8.67 | 11.00 | / | 17.00 |
| The lowest temperature in the cooling process (°C) | 1.06 | -1.39 | 3.90 | 2.12 | 4.11 |
| Snowy days | Once (lasting 2 days) | 2 times (lasting 3 days) | 2 times (lasting 4 days) | Frequent | None |

From October to December 2019, the mean humidity in Wuhan was 75.26% with significantly less precipitation, both of which were much lower than those in the same period in the previous years.

The relatively warm and dry condition may provide favorable meteorological conditions for the survival and reproduction of the virus.

### 2. Meteorological conditions of the onset of SARS pneumonia in 2003

On November 16, 2003, the first case of SARS was reported in Shunde district, Foshan city, and later the infection rapidly spread in Guangzhou city, Guangdong Province. Shunde district is close to Guangzhou city with a 10-km distance, and the climate of two locations are very similar. This explains why the SARS spread to Guangzhou after its onset in Shunde, so we chose Guangzhou city as the subject to conduct a retrospective analysis on the meteorological conditions before (from October 1 to November 15, 2003) and after (from November 16 to December 31) the onset of SARS.

**Table 3** showed that from October 1 to November 15, 2002, the mean temperature of Guangzhou remained above 24 °C and the mean humidity remained at about 73%; from November 16th to December 31st, the temperature of Guangzhou maintained at approximately 16.2 °C and the humidity maintained at approximately 76%. With a relatively stable temperature, SARS in Guangzhou spread from the vector to people and from person to person during this period.

**Table 3.** Comparison of local climate conditions before and after the incidence of the first case of SARS

| Location | Period | Date | Mean temperature (°C) | Mean humidity (%) |
| --- | --- | --- | --- | --- |
| Guangzhou |  | 2002.10.1-10.31 | 23.84 | 72.55 |
|  |  | 2002.11.1-11.30 | 19.39 | 68.17 |
|  |  | 2002.12.1-12.31 | 15.77 | 78.31 |
|  |  | 2003.1.1-1.31 | 13.85 | 69.06 |
|  |  | 2003.2.1-2.28 | 17.83 | 76.52 |
|  | Before | 2002.10.1—11.15 | 23.12 | 69.48 |
|  | After | 2002.11.16-12.31 | 16.18 | 76.33 |
| Beijing |  | 2003.2.1-2.28 | 0.91 | 51.57 |
|  |  | 2003.3.1-3.31 | 6.33 | 56.32 |
|  |  | 2003.4.1-4.30 | 15.29 | 50.23 |
|  |  | 2003.5.1-5.31 | 21.04 | 62.80 |
|  | Before | 2003.3.16-3.31 | 9.59 | 54.44 |
|  |  | 2003.3.16-3.31 (day time) | 14.03 | / |
|  | After | 2003.4.1-4.15 | 13.41 | 49.13 |

On March 15, 2003, the first case of SARS occurred in Beijing. As shown in Table 3, the mean temperature in Beijing in February was only 0.91 °C, which was very unsuitable for the survival of coronavirus, and the mean humidity was 51.57%. The mean temperature in March was 6.33 °C, and the mean temperature during the day was 12 °C. At this time, the low ambient temperature was still not conducive for a large-scale transmission of the coronavirus. By mid-March, the ambient temperature in Beijing further increased. Although the mean temperature of the first half of March was 9.59 °C, the mean temperature in daytime reached 14.03 °C, which was basically suitable for the survival and reproduction of the coronavirus. The mean monthly temperature in April rose to 15.29 °C. During April, the large-scale reproduction of the SARS virus started, and the number of cases of SARS patients kept growing rapidly. The number of patients reached 339 by April 20, and surged to 1,440 by April 30.

### 3. Comparison of meteorological conditions of the onset of 2019-nCoV pneumonia in 2019 and SARS pneumonia in 2003

As shown in **Figure 1**, from October to November 2019 in Wuhan, the mean temperature in Wuhan dropped from 18.28 °C to 13.43 °C. Humidity remained between 73.12% and 77.58%, and the first few cases of the 2019-nCoV pneumonia occurred in early December. The meteorological conditions significantly overlapped with those of SARS onset in Guangzhou. In the winter of 2002, the temperature was between 13.85°C to 15.85 °C, and humidity kept between 69.05% and 78.91% in Guangzhou.

**Figure 1.**
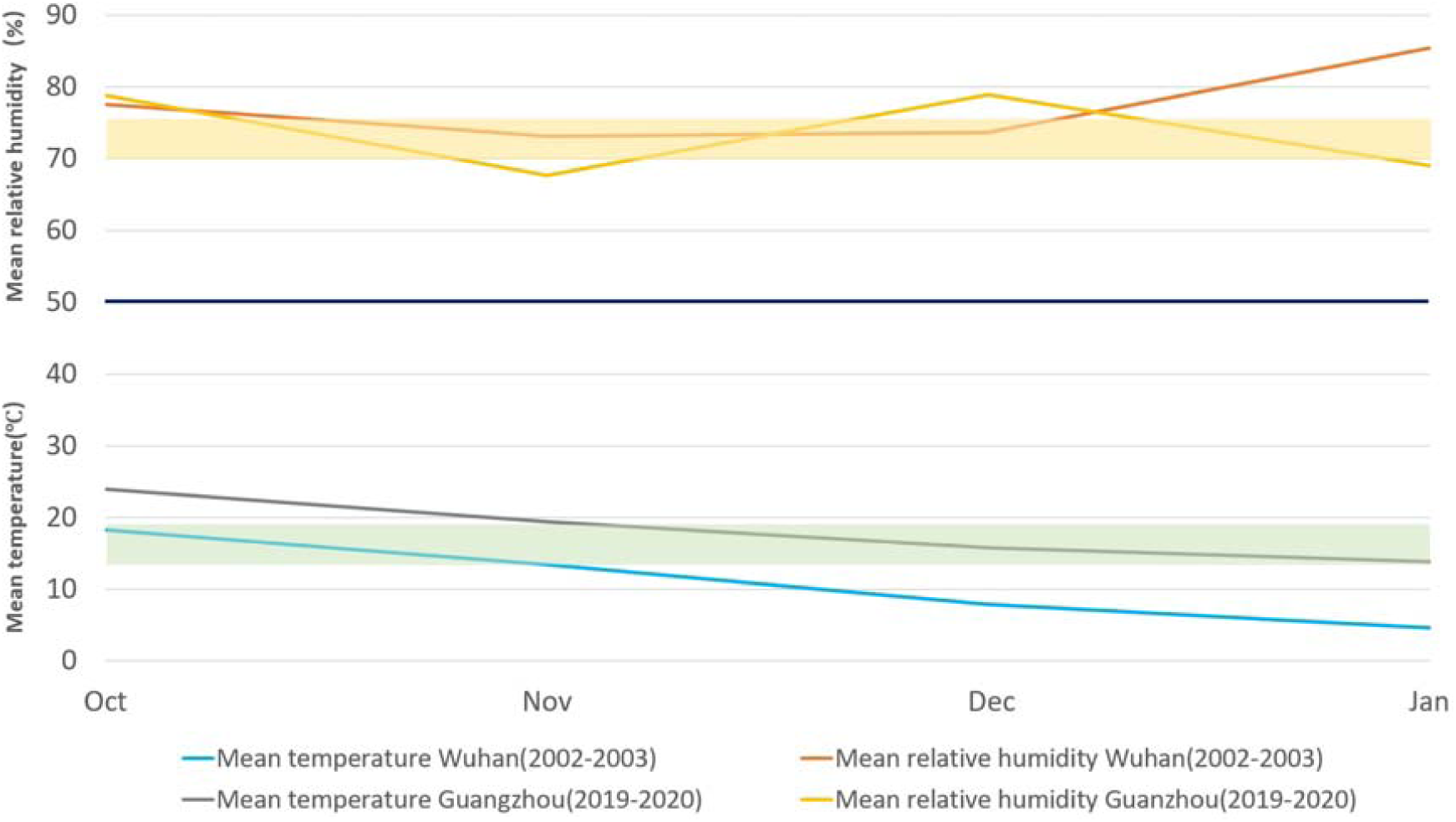
Comparison of meteorological conditions before the onset of 2019-nCoV pneumonia from Oct to Nov, 2019 and SARS pneumonia in Nov to Jan, 2003. From October to November, 2019, the mean temperature in Wuhan dropped from 18.28 °C to 13.4 °C, which was similar to that in Guangzhou between November 2002 and January 2003, during which the temperature ranged from 15.85 °C to 13 °C. The mean humidity of Wuhan was between 73.12% and 77.58%, which overlapped with the humidity interval of 69.06%-78.91% in Guangzhou during 2002-2003.

Considering the data above including 2019-nCoV in Wuhan, SARS coronavirus in Guangzhou and Beijing, we speculate that a meteorological condition with temperature between 13-19°C and humidity between 50% and 80% is suitable for the survival and transmission of the coronavirus.

### 4. Estimation of the possible regression time for the 2019-nCoV infection with reference to SARS data

**Figure 2A** showed that the increase of SARS in Guangzhou entered the platform period at the beginning of May, which means the spread of SARS virus was significantly impressed or stopped, so we chose the mean temperature from April 20 to May 10 (26.26 °C) as an effective temperature for suppressing virus.

**Figure 2.**
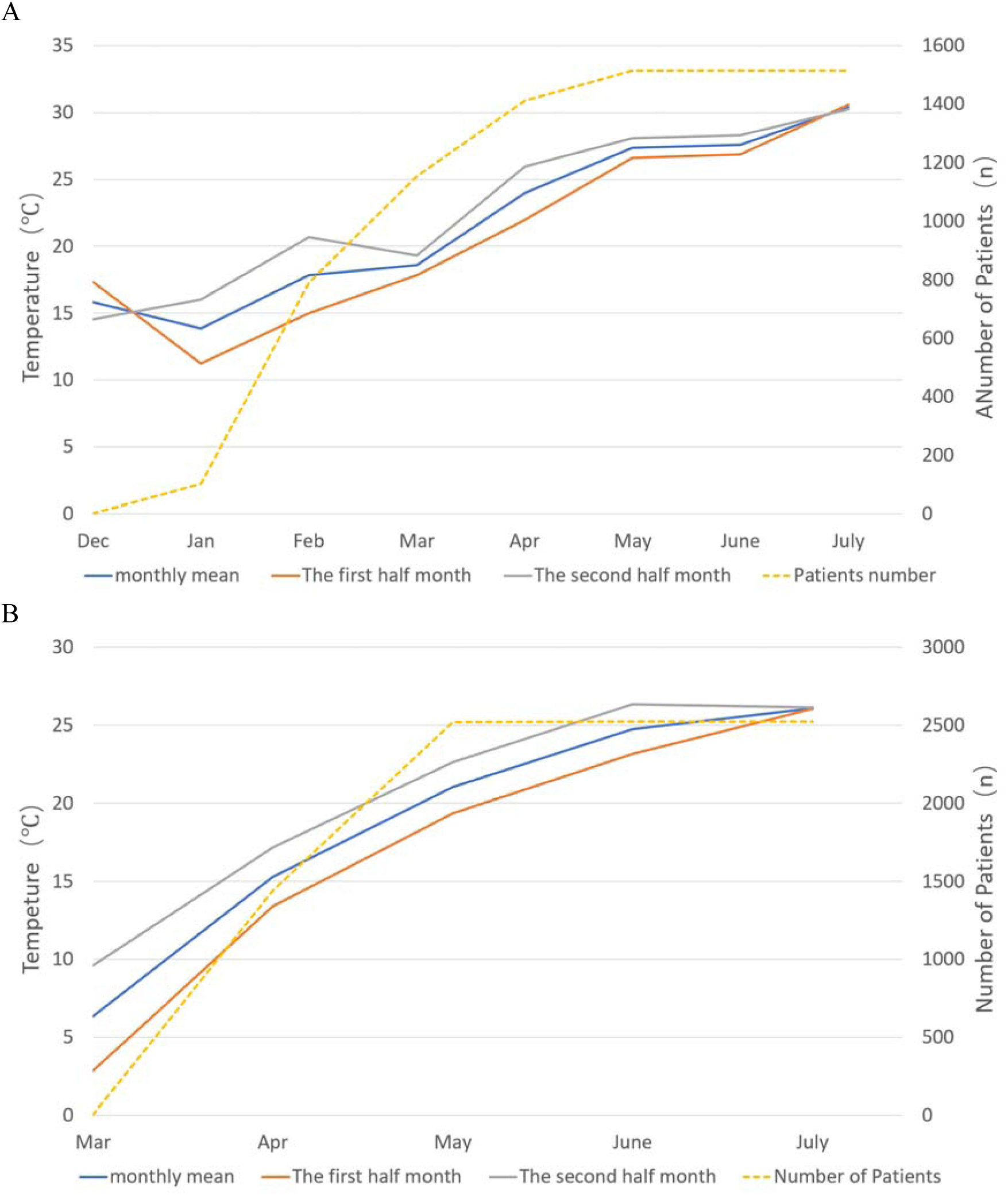
SARS cases and mean temperature changes in Guangzhou (A) and Beijing (B). A: The increase of SARS in Guangzhou entered the platform period at the beginning of May, the mean temperature from April 20 to May 10 (26.26°C) were chosen as an effective temperature for suppressing virus. B: No new case of SARS was reported in Beijing after 20 June, and we chose the mean temperature from June 1 to June 20 (24.22°C) as effective temperature for suppressing virus.

**Figure 2B** showed that by May 30, 2003, the number of cases in Beijing remained at 5,321. During the following 21 days, only two new cases were reported, and no new confirmed cases reported after June 20. It means the spread of SARS virus has completely subsided. We chose the mean temperature from June 1 to June 20 (24.22 °C) as an effective temperature for suppressing virus.

Based on the above data, we found that when the mean temperature reaches 24.22 °C - 26.26°C, the transmission of SARS between patients began to significantly decrease or completely cease. Therefore, with reference to the 2003 SARS virus spreading timeline, the month with a mean temperature of 24.22 °C is expected to be the time of 2019-nCoV pneumonia calming down.

### 5. Global prediction of epidemic trends of 2019-nCoV pneumonia

#### 5.1 Prediction of epidemic trends of 2019-nCoV pneumonia in China

According to the national monthly mean temperature distribution data, with the temperature rises in spring, the 13 °C isotherm gradually moves north. In February, the 13 °C isotherm can reach the southwest of Chongqing, central Hunan, central Jiangxi, and southern Zhejiang. In March, the 13 °C isotherm can reach central Shaanxi, northern Henan, close to southern Hebei. In April, the 13 °C isotherm can reach southern Beijing. In May, apart from the Sichuan Basin, the temperature in other areas can rise to 16 °C or above. The 16-20 °C isotherm reaches central and southern Inner Mongolia, and Beijing will be completely covered by the 24 °C isotherm.

According to the analysis above, in the coming February-April, as the temperature gradually rises from low latitudes to high latitudes, and people move to large urban clusters such as Jiangsu, Zhejiang, Shanghai, Beijing-Tianjin, and the Pearl River Delta, where the mean temperature is between 13 °C to 19 °C, the new coronavirus may break out in these urban agglomerations. **Figure 3A** showed the geographical prediction of infection spread of 2019-nCoV pneumonia in China.

**Figure 3.**
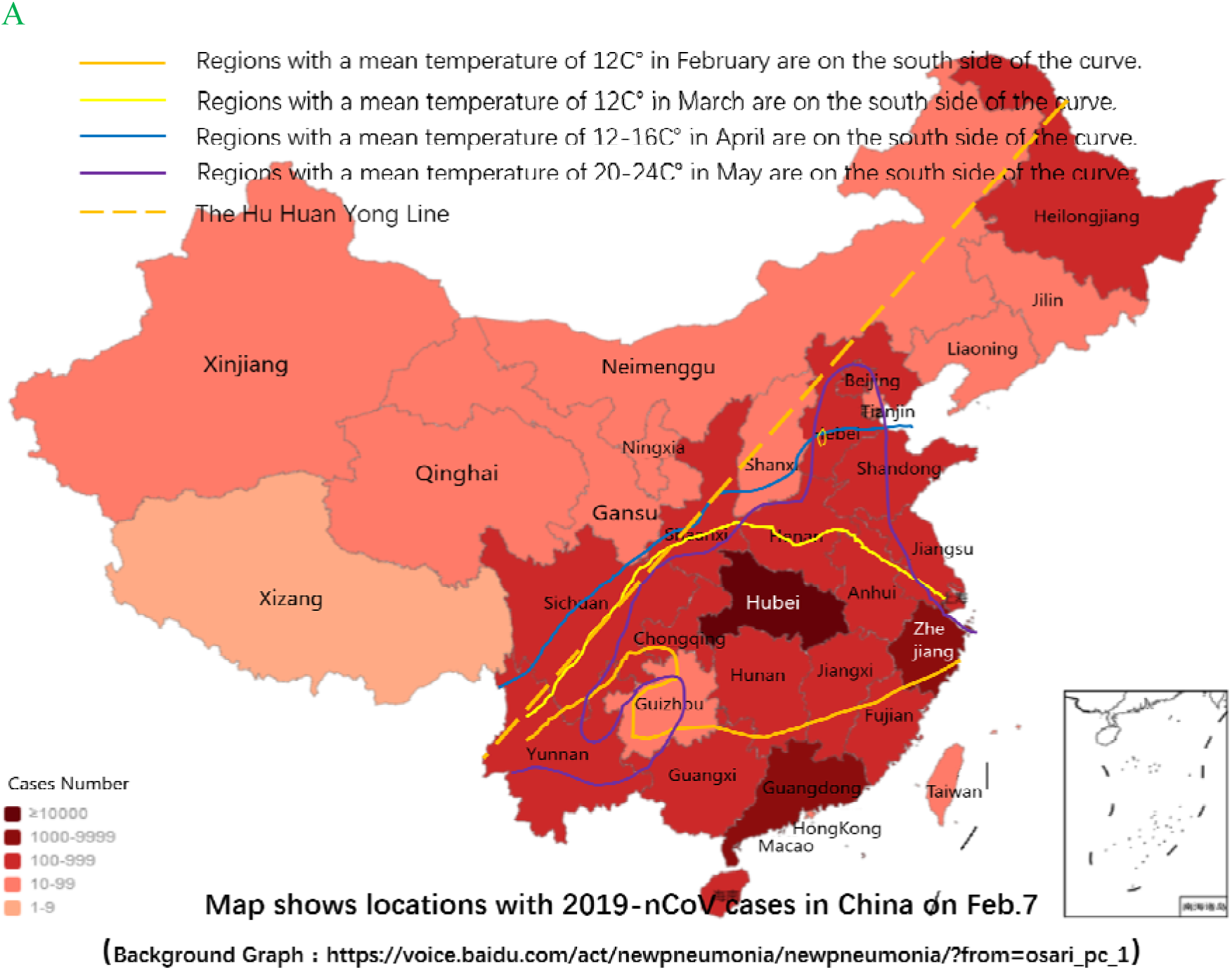
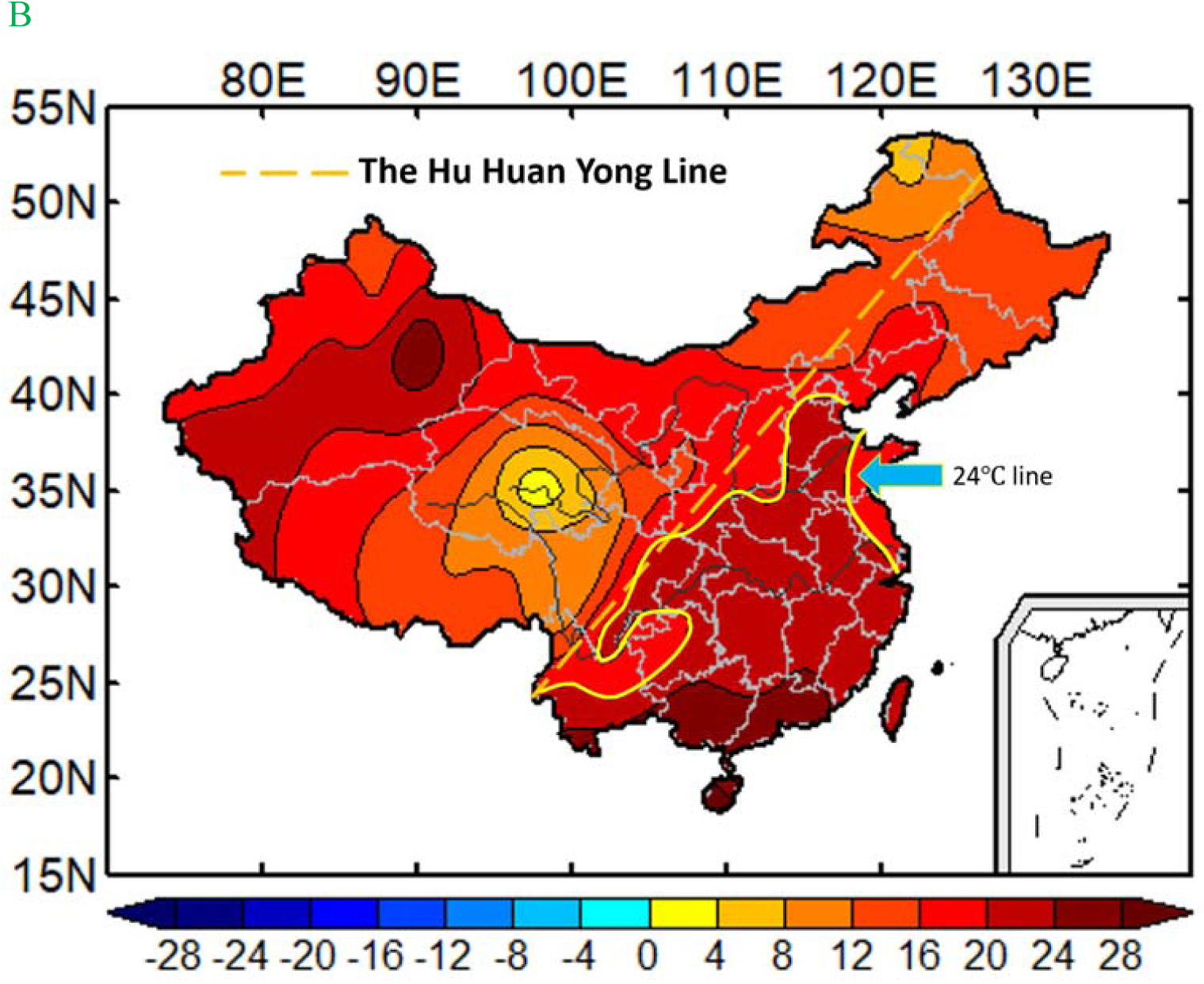
Outbreak trend of 2019-nCoV pneumonia in China. A. Geographical prediction Map of infection development of 2019-nCoV pneumonia in China. B. The mean temperature distribution line in May in China.

**Figure 3B** showed the national mean temperature distribution in May, and we found that the area east of the Hu Huan Yong line, which is the most densely populated region in China, is basically under 24°C climate coverage, and the area is China’s most important urban agglomerations, also the China’s most important population input area, and includes China Beijing-Tianjin-Hebei City Group, Central China Zhengzhou-Wuhan City Group, East China Jiangsu-Zhejiang-Shanghai City Group, and South China Pearl River Delta Group. So, 2020 May is expected to be the time for 2019-nCoV pneumonia to enter a recession period.

#### 5.2 Prediction of epidemic trends of 2019-nCoV pneumonia outside China

From the global mean temperature distribution maps in Figure 4, we can identify the global impact of this 2019-nCoV pneumonia. From February to the end of May, 2020, the areas covered by the temperature (13 °C -24 °C) indicated in the red rectangular are all key areas for disease prevention, especially densely populated cities; from June and thereafter, the disease in regions with a mean temperature over 24 °C will begin to subside.

**Figure 4.**
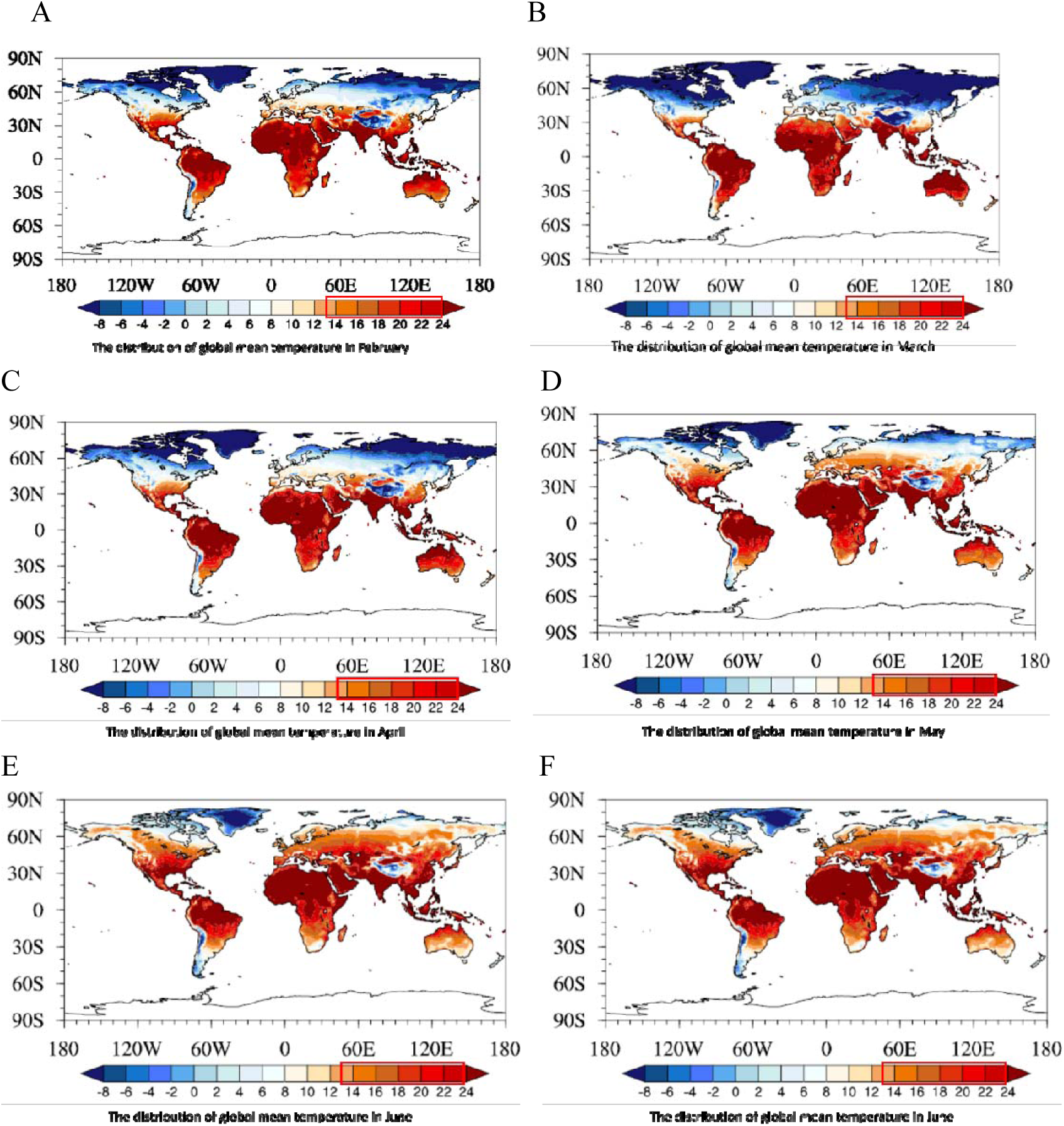
Global impact of this 2019-nCoV pneumonia. The red covers the area under 24 °C. With the coming of spring, the temperature range of 24 °C has gradually expanded in geography. A: World temperature distribution in February; B: World temperature distribution in March; C: World temperature distribution in April; D: World temperature distribution in May; E: World temperature distribution in June; F: World temperature distribution in July.

Prior to June, 2020, cities with a global average temperature below 24 °C are all under high-risk of the transmission of new coronaviruses (**Figure 4**). Aviation departments and ports in various countries need to strictly monitor and prevent the spread of the virus. After June, the risk of disease transmission will be significantly reduced in the cities with a mean temperature reaching 24 °C or higher.

## 6. Discussion

The new coronavirus pneumonia is caused by 2019-nCoV, which is a new pathogen for human being. Due to its outbreak during the Spring Festival travel rush, it spreads at home and abroad in a short period of time. Faced with this new disease, we lack reliable epidemiological information for effective treatment and prevention. However, as we know, any infectious disease origins and spread occur only when affected by certain natural and social factors through acting on the source of infection, the mode of transmission, and the susceptibility of the population. The weather and meteorological factors play a part in coronavirus outbreak besides the social factors^7 8^.

2019-nCoV has a high basic case reproduction number (R0 ranges from 2.2 to 6.7)^9^, causing much more infections than SARS. Because of the biological homology of the coronavirus, we analyzed the meteorological factors of 2019-nCoV outbreak and referenced those of SARS in 2002-2003 to predict the development trend of atypical pneumonia caused by 2019-nCoV and the possible resolution time, which may help the disease control.

Zhou et al^10^ found that coronavirus is probably most active between 9 °C and 24 °C, when the average daily temperature in summer is higher than 24 °C, the possibility of SARS outbreak will decrease. It is consistent with our prediction on the control period of coronavirus. They thought the minimum temperature of coronavirus survival is 9°C, which is lower than 13°C in our study. The difference may come from the Beijing data. In March 2003, the monthly mean temperature was 6.33°C, and the mean temperature from March 16 to March 31 increased to 9.59°C, during which the SARS patients increased. However, by further analysis, we found the mean temperature in day from March 16 to March 31 reached 14.03°C, more closed to that of the onset of SARS in 2002 winter in Guangzhou. April 2003 was the increase period of SARS patient, and it had a monthly mean temperature of 15.29°C, but the day mean temperature from April 1 to April 15 was 13.41°C, a decrease of 1.88°C, so we concluded that the minimum temperature of coronavirus survival is 13°C based on the above analysis.

Warm and dry weather conditions have led to an increase in atmospheric suspended matter, providing conditions for virus adhesion, breeding and transmission. Wuhan had a warm winter of 2019 with a mean temperature of 14.86 °C from October 1 to December 15, which was higher than that of the same period in 2015-2018. Although there were 6 times of cooling event, the cooling process was short with no cooling process being more than 7 days. Because of the short time of cooling, the cold air had no positive effects on the inhibition or killing of virus; on the contrary, it aggravated the infection for the following possible reasons: 1) Cold air can decrease the nasopharyngeal temperature from 36.9°C to 32°C^11^, providing suitable survival and reproduction conditions for coronavirus; 2) Cold air causes vasoconstriction of respiratory tract, further to reduce the immunoglobulin secreted by the mucosa; 3) Dry cold air makes the nasal mucosa prone to small rupture, and creates opportunities for virus invasion^10^. In the same period of previous years, a long period of low temperature played a good role in killing the coronavirus.

Lower rainfall and therefore reduced relative humidity provide a good opportunity for the transmission of respiratory pathogen infections, including coronavirus^4^. Guangzhou had no precipitation for exactly one month before the SARS outbreak at the end of 2002. Precipitation in Wuhan also significantly decreased before the 2019-nCoV outbreak. Although the average monthly humidity of Wuhan from October to December, 2019 was between 69% -77%. The precipitation was significantly reduced to 74 mm, only equivalent to 33.58% of 2015, 37.99% of 2016, and 61.16% of 2018. And the humidity remained at around 50% during the outbreak in Beijing.

Limitation of this study: 1. Due to limited data available, we didn’t include other meteorological factors such as air pressure, atmospheric particles, ultraviolet, and social factors such as population movement for analysis. Inclusion of such factors will provide more accurate and reliable results. 2. As the public data on the new coronary disease and SARS were limited, the numbers of SARS cases in Guangzhou and part in Beijing (before April 20, 2013) were cumulative data of month instead of daily data.

In conclusion, through the analysis of the meteorological conditions for onset of 2019-nCoV and SARS coronavirus, we found that: (1) A wide range of continuous warm and dry weather is conducive to the survival of the virus with a temperature range of 13-24 °C, a humidity range of 50% -80%, a precipitation of 30 mm / month or less. Cold air for more than a week has a significant inhibitory effect on coronavirus. (2) In the coming Feb and April, coronavirus advancing to mid-high latitudes along with an isotherm line of 13-19 °C. The regions with a mean temperature of 13-19°C include the most densely populated areas in the Mainland China, like the Beijing-Tianjin-Hebei area, the Jiangsu-Zhejiang-Shanghai area and other middle-high latitude urban agglomerations. It is therefore necessary to intensify the disease prevention and control in these areas, especially in transportation hub cities. In south China, the Pearl River Delta urban agglomeration should be the focus of disease prevention. (3) The new coronavirus epidemic is expected to end in May, because major domestic urban agglomerations, including the Beijing-Tianjin-Hebei urban agglomeration in central China, the Zhengzhou-Wuhan urban agglomeration in central China, the eastern Jiangsu-Zhejiang-Shanghai urban agglomeration, and the southern Pearl River Delta urban agglomeration, have basically reached 24 °C in May.

## Data Availability

The metereological data, models, or code generated or used during the study are available from the corresponding author (Jing Chen) by request. (List items).

## Acknowledgement

We sincerely thank Jie Guo and Qiao-Mei Wu from Guangdong Meteorological Observation Data Center for their assistance on collection of meteorological data.

## References

1. Wu JT, Leung K, Leung GM. Nowcasting and forecasting the potential domestic and international spread of the 2019-nCoV outbreak originating in Wuhan, China: a modelling study. Lancet 2020 doi: 10.1016/S0140-6736(20)30260-9 [published Online First: 2020/02/06]

2. de Vlas SJ, Cao WC, Richardus JH. Documenting the SARS epidemic in mainland China. Trop Med Int Health 2009;14 Suppl 1:1–3. doi: 10.1111/j.1365-3156.2009.02349.x [published Online First: 2009/10/21]

3. Tan J, Mu L, Huang J, et al. An initial investigation of the association between the SARS outbreak and weather: with the view of the environmental temperature and its variation. J Epidemiol Community Health 2005;59(3):186–92. doi: 10.1136/jech.2004.020180 [published Online First: 2005/02/15]

4. Bi P, Wang J, Hiller JE. Weather: driving force behind the transmission of severe acute respiratory syndrome in China? Intern Med J 2007;37(8):550–4. doi: 10.1111/j.1445-5994.2007.01358.x

5. Cai QC, Lu J, Xu QF, et al. Influence of meteorological factors and air pollution on the outbreak of severe acute respiratory syndrome. Public Health 2007;121(4):258–65. doi: 10.1016/j.puhe.2006.09.023 [published Online First: 2007/02/20]

6. Zhou P, Yang XL, Wang XG, et al. A pneumonia outbreak associated with a new coronavirus of probable bat origin. Nature 2020 doi: 10.1038/s41586-020-2012-7 [published Online First: 2020/02/06]

7. Yip C, Chang WL, Yeung KH, et al. Possible meteorological influence on the severe acute respiratory syndrome (SARS) community outbreak at Amoy Gardens, Hong Kong. J Environ Health 2007;70(3):39–46. [published Online First: 2007/10/19]

8. Yuan J, Yun H, Lan W, et al. A climatologic investigation of the SARS-CoV outbreak in Beijing, China. Am J Infect Control 2006;34(4):234–6. doi: 10.1016/j.ajic.2005.12.006

9. Tian HY. 2019-nCoV: new challenges from coronavirus. Zhonghua Yu Fang Yi Xue Za Zhi 2020;54(0):E001. doi: 10.3760/cma.j.issn.0253-9624.2020.0001 [published Online First: 2020/02/06]

10. Zhou ZX, Jiang CQ. [Effect of environment and occupational hygiene factors of hospital infection on SARS outbreak]. Zhonghua Lao Dong Wei Sheng Zhi Ye Bing Za Zhi 2004;22(4):261–3.

11. Wang J. Health · Environment · Weather Beijing: Meteorological Press 1992:89.

